# Efficacy and safety of tirofiban after intravenous tenecteplase for acute ischemic stroke: the multicenter, randomized, placebo-controlled, double-blind, INSTANT trial protocol

**DOI:** 10.1101/2024.08.28.24312752

**Authors:** Fan Zhang, Minghui Xiao, Cong Zhang, Guangxiong Yuan, Zhiyong Xie, Yi Yin, Ruize Zhou, Shanggui Yuan, Genxiang Xiao, Donghuan Mei, Xiaobing Zeng, Huashi Liu, Huadong Li, Hongwen Liu, Jinchang Tan, Bin Chen, Qingqing Fu, Bin Li, Jinxing Lai, Wei Sun, Shuhua Xie, Zhaohui Lai, Zhongming Qiu, Zidian Jiang, Xianghong Liu, Guoyong Zeng, INSTANT investigators

**Affiliations:** The Affiliated Ganzhou Hospital of Nanchang University, Ganzhou 341000, China; People’s Hospital of Wan’an City, Wan’an 343800, China; Xiangtan Central Hospital, Xiangtan 411100, China; People’s Hospital of Yudu City, Yudu 342300, China; People’s Hospital of Shangyou City, Shangyou 341200, China; People’s Hospital of RuiJin City, RuiJin 342500, China; Second Hospital of Xingguo City, Xingguo 342400, China; People’s Hospital of ShiCheng City, ShiCheng 342700, China; People`s Hospital of County Xunwu, Xunwu 342200, China; People’s Hospital of Anyuan City, Anyuan 342100, China; People’s Hospital of Dayu City, Dayu 341500, China; People’s Hospital of Chongyi City, Chongyi 341300, China; Ganzhou Municipal Hospital, Ganzhou 341000, China; Traditional Chinese Medicine Hospital of Yudu City, Yudu 342300, China; People’s Hospital of Quannan City, Quannan 341800, China; The 903^rd^ Hospital of the People’s Liberation Army, Hangzhou 310000, China; Zhongmeng Hospital of Hexigten Banner, Chifeng City, Hexigten Banner 025350, China

**Keywords:** platelet glycoprotein IIb/IIIa inhibitor, tenecteplase, intravenous thrombolysis, acute ischemic stroke, clinical trial

## Abstract

**Background:** It is very common that symptoms do not improve or even worsen after intravenous thrombolysis (IVT) in acute ischemic stroke. However, it remains unknown whether early administration of tirofiban after IVT may improve clinical outcomes.

**Objective:** This trial aims to assess the efficacy and safety of early administration of tirofiban after intravenous thrombolysis with Tenecteplase in acute ischemic stroke within 24 hours of symptom onset.

**Methods and design:** The INSTANT trial is an investigator-initiated, randomized, placebo-controlled, double-blind, multicenter trial. Up to 310 patients with acute non-large/medium-sized vascular occlusion and non-atrial fibrillation stroke who are treated with intravenous tenecteplase will be consecutively randomized to intravenous tirofiban or placebo in a 1:1 ratio over two years across 50 intravenous thrombolysis-capable stroke centers in China. Subjects will be treated with intravenous tirofiban or placebo at a dose of 0.3 μg per kilogram of body weight per minute for 30 minutes, followed by a continuous infusion of 0.075 μg per kilogram per minute for up to 47.5 hours.

**Outcomes:** The primary efficacy outcome is the proportion of patients with excellent functional outcomes (defined as a score of 0-1 on the modified Rankin scale (mRS)) at 90 days. The secondary outcome is the proportion of patients achieving functional independence, defined as a mRS score of 0-2 at 90 days. Safety outcomes include symptomatic intracranial hemorrhage within 48 hours and mortality at 90 days.

**Discussion:** This randomized, double-blind, placebo-controlled trial will provide data regarding the role of early antiplatelet therapy with intravenous tirofiban after IVT.

**Trial registry number:** ChiCTR2300074368 (www.chictr.org.cn).

## Introduction and rationale

Intravenous thrombolysis (IVT) is currently the first-line medical therapy for acute ischemic stroke (AIS)^1, 2^. However, some studies have found that some patients still suffered severe neurological deterioration after receiving intravenous thrombolysis, which was related to poor 3-month functional outcomes or death^3,4^. Early neurological deterioration (END) within the first 24 hours after IVT for acute ischemic stroke is very common, with the estimates of END incidence varying from 8% to 28%^5^. The reasons for END may related to failure of vascular recanalization, reocclusion, and futile recanalization^6^. Its cause remains unclear, but it may be related to residual fibrinogen and platelet aggregation in microcirculation after IVT^7^. In theory, early antiplatelet therapy may be an effective intervention.

Early administration of antiplatelet therapy may help reduce the risk of END, but it may also increase the risk of symptomatic intracranial hemorrhage (sICH)^6, 8^. Therefore, the effectiveness and safety of antiplatelet therapy within 24 hours after intravenous thrombolysis are still unclear. Tirofiban is a small molecule nonpeptide platelet glycoprotein IIb/IIIa inhibitor (GPI) with pharmacological characteristics, such as non-antigenicity, fast onset effect, short half-life, high selectivity and specificity for platelet glycoprotein IIb/IIIa receptors, and convenient intravenous administration^9^. Tirofiban has been widely used in patients with myocardial infarction^10^; however, it is still an off-label drug for AIS, although it has been applied to some selected patients with AIS in clinical practice.

The SaTIS (Safety of Tirofiban in Acute Ischemic Stroke) trial showed that tirofiban alone did not increase the risk of sICH, any ICH, or mortality^8^. Two studies showed that tirofiban did not significantly reduce mortality rates in patients with AIS^11, 12^. A meta-analysis conducted by Zhou et al. ^13^ showed intravenous tirofiban could be safe for patients with AIS undergoing IVT and increase the likelihood of a favorable functional outcome. Similarly, Yang Q et al. believed that intravenous tirofiban resulted in a greater likelihood of an excellent outcome at 90 days than oral aspirin. Still, only a small proportion of enrolled patients had been treated with IVT^14^. The safety and efficacy of IVT combined with tirofiban remain controversial. Moreover, most of the available data is comprised of using non-standard doses of alteplase for IVT, small sample size, and non-randomized study designs^15^.

Thus, we designed the INtravenouS Tenecteplase ANd Tirofiban for acute ischemic stroke (INSTANT) trial to evaluate the efficacy and safety of early administration of tirofiban after IVT with Tenecteplase in acute ischemic stroke.

## Methods

### Design

The INSTANT trial is a multicenter, double-blind, double-dummy, randomized trial. It is an investigator-initiated trial solely designed in compliance with the Declaration of Helsinki and has been registered at www.chictr.org.cn (identifier ChiCTR2300074368). The protocol has been approved by the ethics committee of Ganzhou People’s Hospital and all participating hospitals before enrollment. The duration of the trial is estimated 2 years. Fifty stroke centers will be included.

### Patient population

#### 2.2.1 Inclusion criteria

1. Age ≥18 years;
2. Patients with no significant improvement (defined as a decrease of 0 or 1 point in the National Institutes of Health Stroke Scale (NIHSS) score compared to baseline) or deterioration (defined as an increase of 4 or more points in the NIHSS score compared to baseline) or fluctuation (defined as an increase of 4 or more points 4 points in the NIHSS score, followed by a decrease of 4 or more points) within 4-24 hours after intravenous tenecteplase for acute ischemic stroke;
3. NIHSS score ≥4 before randomization;
4. Written informed consent is obtained from patients and/or their legal representatives.

#### 2.2.2 Exclusion criteria

1. Intracranial hemorrhage confirmed by CT or MRI after intravenous thrombolysis and before randomization;
2. CTA/MRA/DSA showed occlusion of the internal carotid artery, middle cerebral artery M1 or M2 segment, anterior cerebral artery A1 segment, and vertebrobasilar artery;
3. History of atrial fibrillation;
4. Routine blood test platelet counts less than 100×10^?^/L;
5. Renal insufficiency, glomerular filtration rate < 30 mL/min;
6. Pregnant or lactating women;
7. Allergy to tirofiban;
8. History of neurological or psychiatric illness that precludes the assessment of neurological function;
9. History of bleeding disorder, severe heart, liver or kidney disease, or sepsis;
10. CT or MR evidence of mass effect or intracranial tumor (except small meningioma);
11. CT or MR angiography evidence of intracranial arteriovenous malformations or aneurysm;
12. Any terminal illness with a life expectancy of less than 6 months;
13. Participating in other clinical trials.

##### Randomization

Randomization will be performed immediately via a web-based App on a mobile phone or computer (https://jinlingshu.com) after the patient’s eligibility has been established. Eligible patients will be randomly allocated in a 1:1 ratio to tirofiban or control group. All trial personnel and patients will be unaware of the treatment assignment.

##### Contents of study drug kit

Tirofiban and its placebo (saline) are manufactured and provided by Lunan Pharmaceutical Group Co., Ltd., Linyi, China. The medication bottles are visually identical (including labeling, dosage form, size, and color) except for a unique number.

#### 2.4.1 Tirofiban group

The box of the tirofiban group contains a tirofiban hydrochloride injection (5mg/100ml * 5 bottles; the bottles are marked as Study Drug), aspirin, and clopidogrel tablet placebo (one piece each).

#### 2.4.2 Control group

The box of the control group contains saline placebo (100ml * 5 bottles, marked as Study Drug), aspirin, and clopidogrel tablet (one piece each).

##### Treatments

All patients are treated with intravenous tenecteplase first, and patients who meet the selection criteria will be randomly assigned to either the tirofiban or placebo group in a 1:1 ratio. Patients assigned to the tirofiban group will be treated with intravenous tirofiban. Patients in the placebo group will not be treated with intravenous tirofiban. The relevant time points (to the minute) and treatment information must be recorded accurately.

#### 2.5.1 Tirofiban group

Patients assigned to the tirofiban group will be treated with intravenous tirofiban. It is recommended to use tirofiban as soon as possible (within 10 minutes recommended) after randomization. Intravenous tirofiban will be administered at a dose of 0.3 μg per kilogram of body weight per minute for 30 minutes, followed by a continuous infusion of 0.075 μg per kilogram per minute for up to 47.5 hours. Aspirin placebo (1 tablet) and/or clopidogrel placebo (1 tablet) will be given orally at 24 hours after intravenous tenecteplase. Antiplatelet therapy with aspirin (100 mg) and/or clopidogrel (75 mg) will be administered at 44 hours after randomization until the follow-up period of 90 days.

**Table 1.** Visit plan and data collection.

|  | Baseline | Day 1~2 | Day 5~7 | Day 90 |
| --- | --- | --- | --- | --- |
| Eligibility criteria | X |  |  |  |
| Informed consent | X |  |  |  |
| Randomization | X |  |  |  |
| Demographics | X |  |  |  |
| Medical history | X |  |  |  |
| Clinical examination | X | X | X |  |
| mRS score | X |  |  | X |
| NIHSS score | X | X | X |  |
| Laboratory results | X |  |  |  |
| Electrocardiography | X |  |  |  |
| Brain CT+CTA or MRI+MRA | X | X |  |  |
| Concomitant medication |  | X | X | X |
| Adverse event |  | X | X | X |
| EQ-5D-5L |  |  |  | X |

Acronyms: ASPECTS denotes Alberta Stroke Program Early CT Score, CTA CT angiography, EQ-5D-5L European Quality Five Dimensions Five Level scale, MRA MR angiography, mRS modified Rankin Scale, and NIHSS National Institutes of Health Stroke Scale.

#### 2.5.2 Placebo group

Patients in this group will receive intravenous placebo (saline), not tirofiban treatment. It is recommended to use a placebo as soon as possible (within 10 minutes recommended) after randomization. Aspirin (100 mg) and/or clopidogrel (75 mg) will be given orally at 24 hours after intravenous tenecteplase. Antiplatelet therapy with aspirin (100 mg) and/or clopidogrel (75 mg) will be administered at 44 hours after randomization until the follow-up period of 90 days.

The treatment scheme is shown in Figure 1.

**Figure 1.**
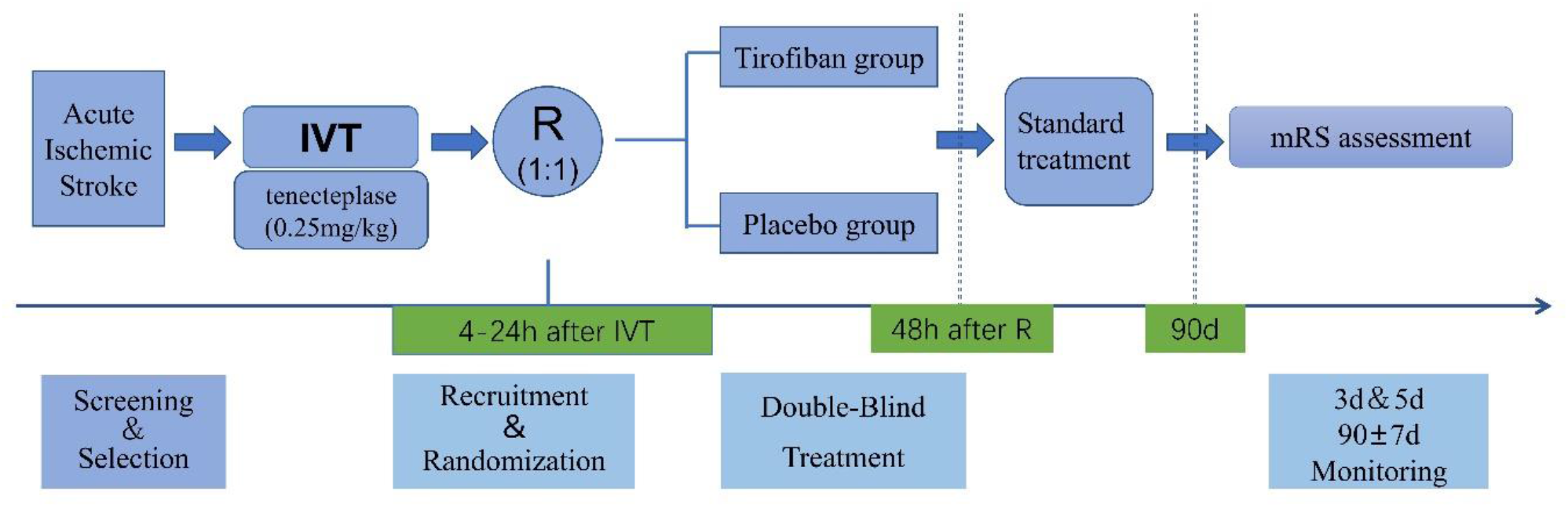
Trial design and treatment flow diagram. Shown are the study design and treatment flow diagram of the INSTANT trial. Acronyms: mRS modified Rankin Scale, IVT intravenous thrombolysis, and R randomization.

However, the use of any other (intravenous or oral) anticoagulants or antiplatelet agents is not allowed after randomization to the 90-day follow-up period.

### Efficacy end-points

#### 2.6.1 Primary end-point

The primary efficacy endpoint is an excellent outcome, defined as a score of 0 or 1 on the modified Rankin scale (mRS) at 90 days after randomization. The mRS scores will be centrally adjudicated by two certified neurologists based on video or voice recordings at 90-day follow-up. For those who decline to participate in a video or voice recording, the outcomes will be determined in person by site neurologists blinded to the treatment allocation.

#### 2.6.2 Secondary efficacy end-points

1. mRS score at 90 days (shift analysis);
2. The proportion of patients functionally independent (mRS score 0 to 2) at 90 days;
3. The proportion of patients ambulatory or bodily needs capable or better (mRS score 0 to 3);
4. Early neurologic improvement, defined as the NIHSS score at 48 hours after randomization, is reduced by 30% or more compared to the NIHSS score at randomization;
5. Health-related quality of life, assessed with the European Quality Five Dimensions Five Level scale (EQ-5D-5L) at 90 days.

#### 2.6.3 Safety end-points

1. Symptomatic intracranial hemorrhage (SICH) defined as per the Heidelberg bleeding classification;
2. Radiologic intracranial hemorrhage rate within 48 hours;
3. Mortality at 90 days;
4. Incidence of non-hemorrhagic serious adverse events, such as pneumonia, respiratory failure, circulatory failure, cerebral herniation, secondary epilepsy, sepsis, renal failure, acute coronary syndrome, venous thrombosis, etc;
5. Other serious adverse events.

##### Data and Safety Monitoring Board (DSMB)

The independent DSMB will consist of 3 experts in the fields of stroke and biostatistics. All of them are neither trial participants nor affiliated with the study sponsors. The DSMB will meet annually and monitor trial progress. In addition, the DSMB will review the incidence of serious adverse events to ensure patients’ safety.

##### Sample size estimates

According to an observational study conducted in China, we assumed that the proportion of excellent outcomes in the tirofiban and placebo groups is 38% and 23%, respectively^16^. Based on these data, taking an attrition rate of 5% into account, we estimated that 155 subjects in each group (310 in total) will provide 80% power to detect a difference of 15% in the excellent outcome between the two groups at the two-sided 0.05 significance level. This study will have an interim analysis of SICH, which is not an excellent outcome. This estimation is performed based on PASS (NCSS, LLC. Kaysville, Utah, USA) version 15.0.

##### Statistical analyses

The primary analysis of the INSTANT trial will compare the 90-day mRS 0-1 proportion between the two treatment groups. The effect variable of primary outcome is the odds ratio and will be estimated with binary logistic regression. The secondary outcomes and safety outcomes will be analyzed using χ^2^ tests, t-tests, Mann-Whitney U tests, and multivariable linear, ordinal, or binary logistic regression as appropriate. The adjusted odds ratio, common odds ratio, and beta coefficient will be adjusted for the following main prognostic variables: age, baseline NIHSS score, and time from onset to randomization. Treatment effect modification will be evaluated in subgroup analyses based on the above variables and other variables of interest. All statistical analysis will be conducted based on the intention-to-treat population with the per-protocol population as a reference. Patients who actually receive the allocated treatment and do not have major protocol violations will be included in the per-protocol analysis. The missing data of baseline variables will be imputed with multiple imputations. Missing outcome data of mRS scores at 90 days in the lead analysis will be imputed using multiple imputations. Complete case and worst-case analyses will be performed as sensitivity analyses^17^. In the worst-case analysis, if the patient was identified as alive, we will assign a score of 5. Otherwise, we will impute a score of 6. Statistical analysis will be performed on the SAS 9.4 system.

## Discussion and summary

Early neurological deterioration (END) within the first 24 hours after IVT for acute ischemic stroke is comparably common; unfortunately, there is a lack of effective treatment measures. Previous studies have suggested that intravenous tirofiban is a potentially effective treatment strategy^18^. The INSTANT trial is designed to investigate the efficacy and safety of early administration of tirofiban within 24 hours of symptom onset after intravenous tenecteplase in acute ischemic stroke. Our trial randomized first patients in April 2024 and the duration of the trial is estimated 2 years. Excellent outcome is the primary endpoint of INSTANT and is a widely accepted endpoint in this population.

The RESCUE BT trial suggests that tirofiban may increase the risk of intracranial hemorrhage in patients with cardiogenic stroke^18^. The pathogenesis mechanism of cardiogenic stroke manifested as a clinical syndrome in which emboli from the heart and aortic arch pass through the circulation and cause cerebral artery embolism and corresponding brain dysfunction, rather than platelet activation, aggregation, and adhesion^19^. Therefore, this study excluded patients with cardiogenic stroke, such as atrial fibrillation. The INSTANT randomized trial will provide important data on the effectiveness and safety of antiplatelet therapy within 24 hours after intravenous thrombolysis due to acute ischemic stroke patients presenting within 4.5 hours of last known well. This trial will be conducted concurrently with the RESCUE-BT3 trial(ChiCTR2400080653) led by Qingwu Yang et al., the ADVENT trial (NCT05199194) led by Yi Yang et al. ^20^. and the ASSET-IT trial (NCT06134622) led by Wei Hu et al. in China.

We look forward to merging the data after completing these four trials and conducting meta-analysis at both the study and individual levels. The INSTANT study had several limitations. All participants in this trial were patients who had undergone intravenous thrombolysis treatment. In order to minimize the risk of intracranial hemorrhage, the trial steering committee and the principal investigator decided to reduce the dose of tirofiban by 1/4 according to the RESCUE-BT2 trial ^21^ after consultation. However, further research is needed to determine the optimal dosage of tirofiban in this population. Second, this trial excluded patients with large vessel occlusion stroke, mild stroke (NIHSS score<4), thromboembolic stroke, as well as patients who underwent endovascular thrombectomy, which may limit the generalizability of the results. In addition, we only enrolled patients who received intravenous tenecteplase; therefore, the results may not be generalizable to patients who were treated with intravenous alteplase. The INSTANT randomized trial will provide important data on the effectiveness and safety of antiplatelet therapy within 24 hours after intravenous thrombolysis due to acute ischemic stroke patients presenting within 4.5 hours of last known well.

## Data Availability

not applicable

## Non-standard Abbreviations and Acronyms

IVT: intravenous thrombolysis
mRS: modified Rankin scale
AIS: acute ischemic stroke
END: early neurological deterioration
sICH: symptomatic intracranial hemorrhage
GPI: glycoprotein IIb/IIIa inhibitor
NIHSS: National Institutes of Health Stroke Scale

